# Evaluation of Different Types of Face Masks to Limit the Spread of SARS-CoV-2 – A Modeling Study

**DOI:** 10.1101/2021.04.21.21255889

**Authors:** Brian M. Gurbaxani, Andrew N. Hill, Prabasaj Paul, Pragati V. Prasad, Rachel B. Slayton

## Abstract

We updated a published mathematical model of SARS-CoV-2 transmission with laboratory-derived source and wearer protection efficacy estimates for a variety of face masks to estimate their impact on COVID-19 incidence and related mortality in the United States. When used at already-observed population rates of 80% for those ≥65 years and 60% for those <65 years, face masks are associated with 69% (cloth) to 78% (medical procedure mask) reductions in cumulative COVID-19 infections and 82% (cloth) to 87% (medical procedure mask) reductions in related deaths over a 6-month timeline in the model, assuming a basic reproductive number of 2.5. If cloth or medical procedure masks’ source control and wearer protection efficacies are boosted about 30% each to 84% and 60% by cloth over medical procedure masking, fitters, or braces, the COVID-19 basic reproductive number of 2.5 could be reduced to an effective reproductive number ≤ 1.0, and from 6.0 to 2.3 for a variant of concern similar to delta (B.1.617.2).

**Article Summary Line:** Adapting a published SARS-CoV-2 transmission model together with updated, laboratory-derived source control and wearer protection efficacy estimates for a variety of face coverings as well as N95 respirators, we demonstrate that community masking as currently practiced has likely reduced cases and deaths and that this benefit can be increased with wider adoption of better performing masks.

## Introduction

The emergence of coronavirus disease 2019 (COVID-19) has had a substantial impact on populations globally, with efforts across governments to prevent its remarkable spread. While social distancing has been universally recommended since very early in the pandemic, recommendations for masks in the general population were adopted later in many countries (see, for example, [1]). Several factors contributed to the initial uncertainty around the potential impact of widespread use of face masks on SARS-CoV-2 transmission. A large and well-designed 2015 study on cloth face masks (the main type of mask available to the public at the time) contributed to the scientific uncertainty that these types of face coverings were effective for preventing the transmission of respiratory diseases [2]. There were initial hypotheses that cloth masks could give the wearer a false sense of protection and even contaminate the wearer with accumulated viral particles, notably described in a high-profile study in the *Annals of Internal Medicine* that was later retracted (for failure to note PCR assay values that were below the limit of detection) [3]. Furthermore, a major concern at the beginning of the outbreak in the US was supply, especially of high-quality masks like N95 respirators. As it became clear, however, that the virus can spread through exhaled respiratory droplets from infected individuals without symptoms [4], the U.S. Centers for Disease Control and Prevention (CDC) recommended masks for general use early in the U.S. pandemic (as of April 2020, [5]). Evidence continues to show that asymptomatic and clinically mild infections contribute substantially to SARS-CoV-2 transmission [6-9]. Together, this growing body of evidence has highlighted the importance of prevention measures, like masking, to reduce transmission from people who are asymptomatic, undetected, or both.[6-8].

As the COVID-19 pandemic has continued, evidence has accumulated that face mask use by the general population can limit the spread of SARS-CoV-2. This evidence has taken three main forms, described in order of their appearance in the literature: 1) modeling studies that suggested that even if masks are limited in their efficacy, widespread use across the population could still reduce the spread of the virus to a considerable degree [10, 11], 2) laboratory studies that demonstrated masks physically block exhaled droplets and aerosols containing virus from infected persons (source control) and also offer wearer protection [12-14], and 3) epidemiological studies that documented lower transmission in settings where masks were used [15-19]. In this study, we extend the model of Worby and Chang to use age-stratified social contact patterns for the general U.S. population, and we analyzed the model both employing the measured face mask efficacy parameters for a variety of specific types of masks and for efficacy estimates that can act as benchmarks for evaluating these products [20].

## Methods

We adapted the transmission model (used for studying resource allocation of masks) of Worby and Chang (2020) for face mask adoption in a hypothetical population by expanding it to the age-stratified social contact patterns characteristic of the demographic profile of the United States. The underlying structure of this compartmental model is described in Worby and Chang [20], which we briefly summarize. Individuals are classified according to their disease status and whether or not they wear a mask in public. The model is further stratified by age in 5-year age bands. People contact each other (defined as either direct physical contact, e.g. through a handshake or a kiss, or a proximal, two-way conversation of 3 or more words) at age-specific daily rates estimated for the United States, as described by Mossong et al. and Prem et al. [21, 22]. We compared the results of the model with the age stratification removed, and the results were significantly different (data not shown). Given that the infection fatality ratios (IFRs) are strongly age structured, we believe the age stratification is appropriate. Vaccination is not explicitly part of the model and has not been included in this study.

A schematic of the compartmental model is shown in Figure 1. Susceptible individuals who are infected move into an exposed compartment and thereafter into a pre-symptomatic compartment. Subsequently, a pre-specified proportion of these individuals moves into an asymptomatic state, while the remainder become fully symptomatic. Pre-symptomatic, asymptomatic, and fully symptomatic SARS-CoV-2 infected individuals all contribute to the force of infection with varying degrees of infectiousness. All asymptomatic individuals recover, whereas a proportion of fully symptomatic individuals do not recover and die. A fraction of asymptomatic cases is assumed to be detected whereupon a fraction of these individuals begins to use a mask and continue to mask thereafter. Similarly, a fraction of symptomatic cases is assumed to know they have COVID-19, and these individuals put on a mask at the same adoption level as detected in asymptomatic cases. Symptomatic persons and detected, asymptomatic persons who wear a mask also change their contact rates reflective of some degree of isolation/quarantine. We do not include specific compartments modeling quarantine per se, but rather we reduce contact rates which accomplishes the same purpose and maintains simplicity of compartmental structure while allowing a degree of mixing that might be anticipated among a fraction of infected individuals who are not strictly isolating themselves. We also assumed a fraction of the general population adopts mask usage at the outset and continues usage regardless of infection status. Other than the aforementioned masked cases, we assumed that contact rates among age groups remain the same when people wear a mask. A basic reproduction number (R_0_) of 2.5 was assumed in the absence of any mask use, consistent with CDC’s pandemic planning scenarios [23]. We also explored the model with a basic reproduction number of 4.0, in keeping with the estimated magnitude of the B.1.1.7 variant [24]. The modeled time horizon was 6 months and the cumulative number of infections and deaths were recorded. The impact of various levels of mask adoption was assessed by calculating the relative reduction in cumulative infection and deaths, comparing cumulative cases and deaths to the same model over the same time horizon with no mask use in the entire population.

**Figure 1:**
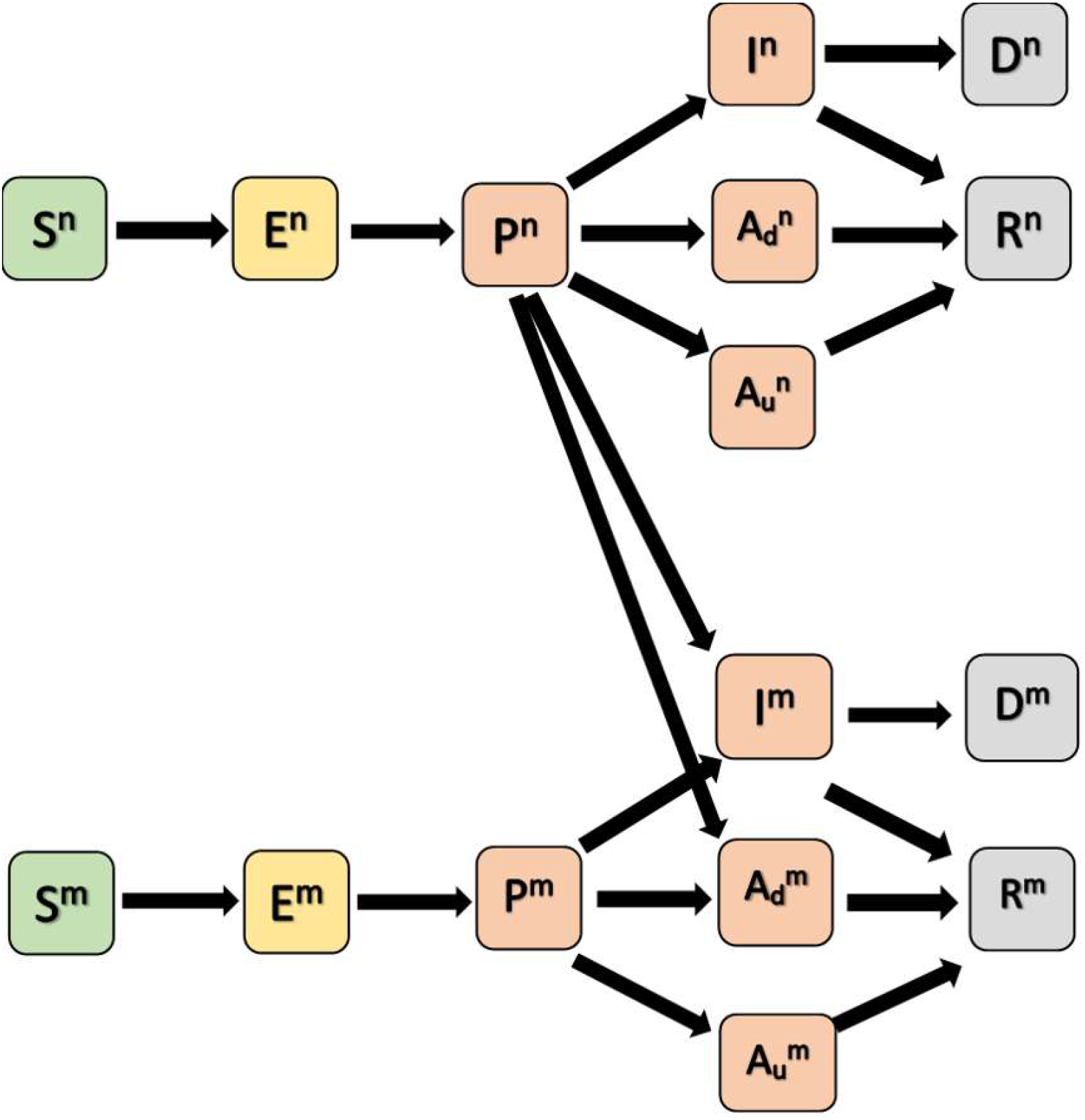
Schematic of compartmental model. Compartments are susceptible (S, green), exposed (E, yellow), infectious compartments (pre-symptomatic P, asymptomatic and detected A_d_, asymptomatic and undetected A_u_, symptomatic I, pink), recovered (R, gray), and died (D, gray). Superscript ‘n’ denotes no mask, and ‘m’ denotes mask.

Masks were modeled to reduce transmission via two different mechanisms: source control efficacy, whereby mask wearing by an infectious person reduces their likelihood of transmitting SARS-CoV-2; and wearer protection efficacy, whereby masks protect a susceptible person from becoming infected when exposed to an infectious person. We examined adoption of various kinds of masks (e.g., cloth, medical procedure, N95 respirators) specifically incorporating estimates from a recent study of source control efficacy [14]. A range of values of hypothetical wearer protection efficacy was assumed for each kind of mask. Although it has generally been found that wearer protectiveness coefficients are approximately half the source control values [13, 25, 26], wearer protection efficacy was allowed to vary in the plot because it could be greatly affected by how the mask is worn, maintained, and used. Characteristics of each mask when worn according to manufacturers’ specifications can be found in Lindsley et al. and are shown in Table 1 [14]. We do not address the issue of mask and respirator use in healthcare settings in this paper, as there is substantial public health guidance regarding the use of personal protective equipment in healthcare settings [27].

**Table 1:** Parameter values used in the simulation

| Parameter | Value [reference] |
| --- | --- |
| Percentage of uninfected persons wearing masks at the outset | Varies by scenario |
| Mask efficacy as source control: | [14] |
| N95 respirator | 96% |
| Medical procedure mask | 56% |
| Cloth mask | 49% |
| Gaiter | 48% |
| Bandana | 33% |
| Percentage of asymptomatic detected and symptomatic detected COVID-19 cases who adopt mask use | <65 years old: 70%<br>≥65 years old: 90% |
| Percentage of symptomatic cases who know they have COVID-19 | 18.3% (see Supplement) |
| Average duration of incubation period – other SARS-CoV-2 | 6 days [23] |
| – Delta VOC | 4 days [51, 52] |
| Average duration of asymptomatic and symptomatic periods | 9 days [23] |
| Relative infectiousness of asymptomatic cases/symptomatic cases | 75% [23] |
| Percentage of infections that are asymptomatic | 30% [23] |
| Probability of detecting asymptomatic case | 10.7% (see Supplement) |
| Infection Fatality Ratio (IFR)<br>(IFR's for Delta VOC $\approx 2x$ shown for other SARS-CoV-2)<br>[53, 54] | Ages 0–19: 0.00003<br>Ages 20–49: 0.0002<br>Ages 50–69: 0.005<br>Ages 70+: 0.054 [23] |
| Reduction in contact rate for symptomatic and detected<br>asymptomatic persons wearing a mask | 50% |
| Risk of death for symptomatic cases | See Supplement for<br>calculation |

## Results

Figure 2 depicts heat maps of reduced transmission and deaths over 6 months as a function of varied source control efficacy and wearer protection efficacy. Mask wearing rates by the various sub-populations in the model are provided in the figure caption. These rates were in line with surveys of mask usage in the United States in May and June 2020 [28]. The colored bands of the plots represent contours of relative reduction. Going from the bottom left corner of the figures (source control efficacy and wearer protection efficacy both 0%, equivalent to no mask wearing in the population) these increase in 5% increments to the right top corner (source control efficacy and wearer protection efficacy both 100%). For example, to obtain at least a 50% reduction in cumulative infections, source control would need to be at least 55% efficacious in limiting transmission in the population for arbitrary wearer protection efficacy. Source control would need to be approximately 45% effective to reduce the number of deaths by half regardless of wearer protection efficacy.

**Figure 2:**
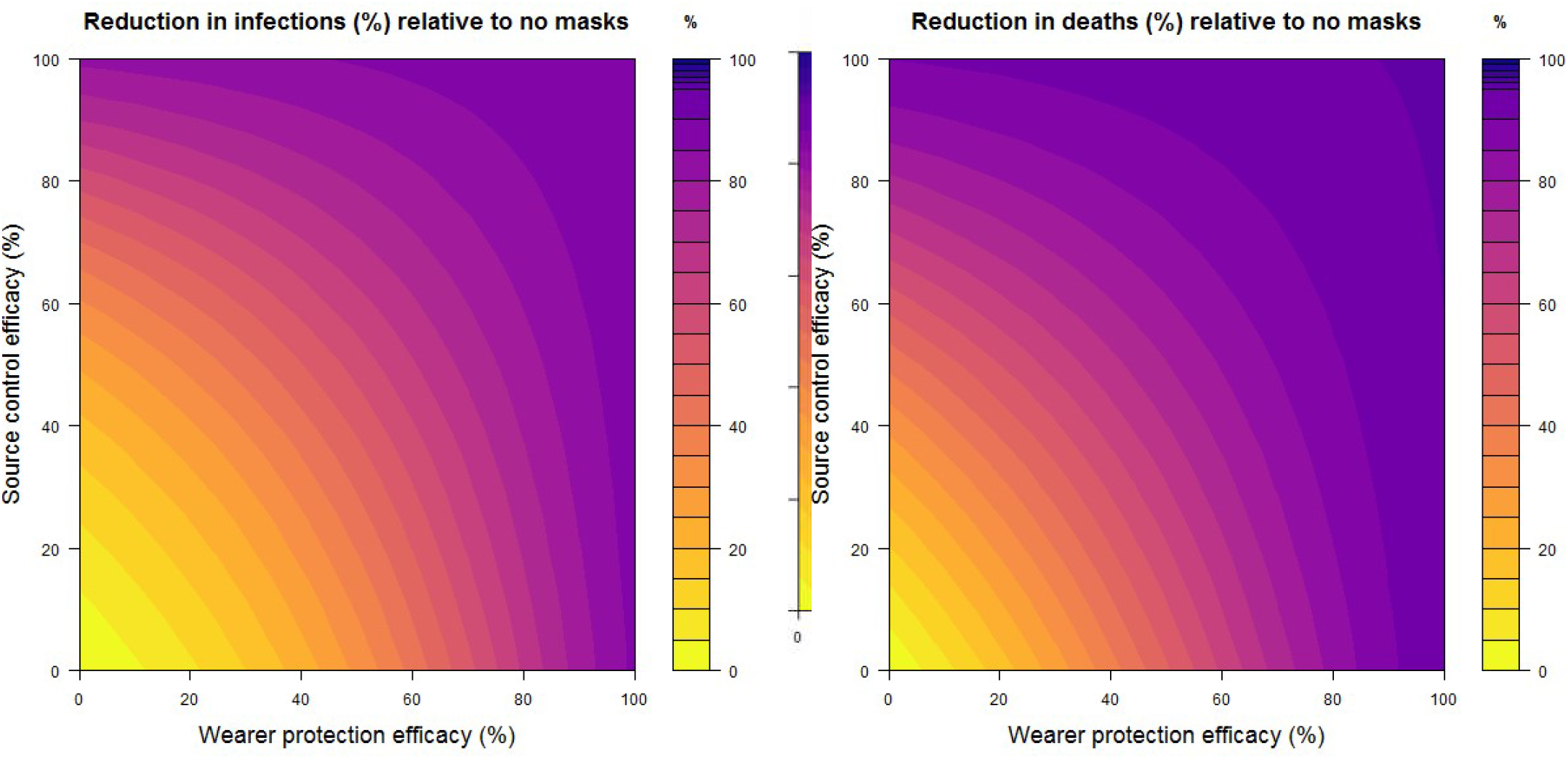
Heat maps of the percentage reduction in cumulative infections at the end of 1 year relative to no mask use in the population, assuming a baseline R0 = 2.5. Assumes 60% of the susceptible population <65 years old are wearing masks, 80% of those ≥65 years old wear masks, and both rates increase 10% for detectably infected persons (whether symptomatic or asymptomatic). The simulation posits that 18.3% of symptomatic infected people and 10.7% of asymptomatic infected individuals have been detected by screening and are known to be carrying SARS-CoV-2 (see the Supplement). Mask efficacy parameters for source control and wearer protection increase along the vertical and horizontal axes, respectively. Reductions in cumulative infections over 6 months are shown on the left; reductions in deaths are shown on the right.

Even with the source control and wearer protection efficacy for the types of mask that most wearers are likely to use, such as medical procedure or cloth masks and gaiters (see Table 1), substantial reductions in case load and death can be achieved with general population use at stated levels. Even at lower levels of use, reductions are estimated to be substantial As source control and wearer protection efficacy approach 100% for the masks, relative reduction in infections also approaches 100%, even though mask adherence is far from 100%, because transmission dips below the epidemic threshold (i.e. an effective reproduction number < 1). Our simulations project that a 70% reduction in cumulative infections, relative to zero mask usage, could be achieved with hypothetical combinations of wearer protection and source control efficacies, respectively, of (0%, 65%), (25%, 50%), (40%, 35%), (50%, 25%), among many others lying on the 70% contour curve of the left panel of Figure 2.

Figure 3 depicts the reduction in infections with different population-wide percentages of mask use, with the assumption that mask wearer protection efficacy is half of source control efficacy and that mask use among persons <65 years old is 70% that of persons ≥65 years old. We evaluated these impacts for SARS-CoV-2 (3A, left) and one of its highly contagious variants of concern (3B, right, for parameters similar to the Delta variant). Mask wearing rates for detected and infected people are fixed at 90% for those ≥65 years old, and 70% for those who are younger. Based on the model, in Figure 3A if 25% of the general population ≥65 years old puts on a mask, cumulative cases after 6 months are reduced by 23% (N95), 14% (medical procedure), 12% (cloth), 12% (gaiter), and 9% (bandana). If mask adoption is 50% for the general population ≥65 years old, projected reductions in cases are 57% (N95), 32% (medical procedure), 28% (cloth), 28% (gaiter), and 20% (bandana). If mask adoption is 75% for ≥65 years old, projected reductions in cases are 95% (N95), 65% (medical procedure), 55% (cloth), 54% (gaiter), and 35% (bandana). Note that even with 0% mask use for the susceptible population (horizontal axis), there is still a significant measure of infection control because of mask adoption among detected infected people.

**Figure 3:**
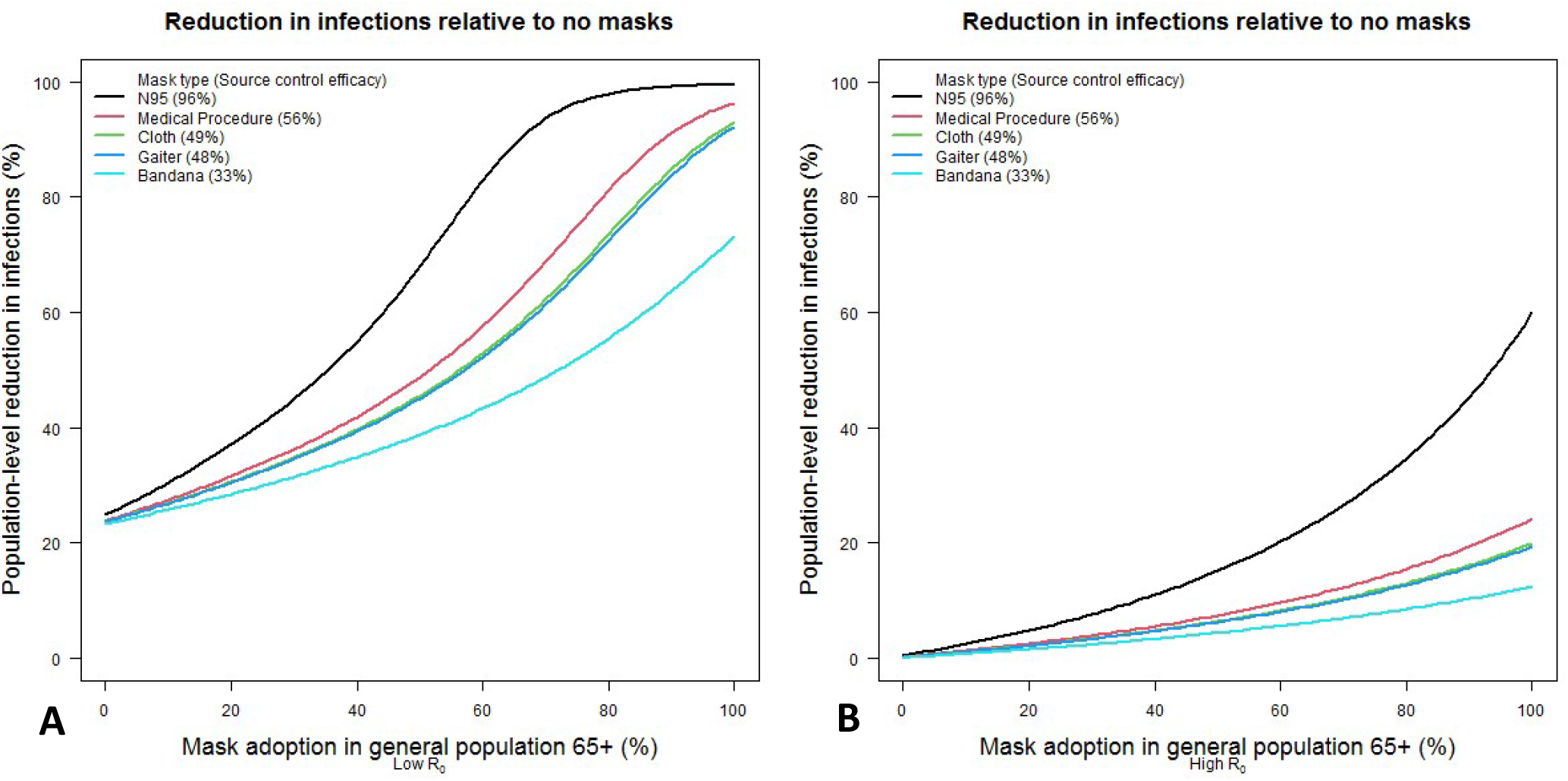
The percentage reduction in cumulative infections after 6 months of simulation, relative to no mask use in the population, as mask use varies in the general, susceptible population for different types of face masks. Mask source control parameters are fixed according to estimates for the given types, and wearer protection efficiency is assumed to be half of source control effectiveness. In this analysis, younger susceptible persons are assumed to use masks at 70% of the rate of persons ≥65 years old. Known infected people ≥65 years old are masked at a 90% rate, with younger persons at 70%. The baseline R_0_ in the absence of mask use is assumed to be 2.5 in the left panel and 6.0 in the right panel.

Figure 3B shows similar results to 3A, but assuming a much more highly contagious variant, similar to Delta (B.1.617.2) with an R0 = 6.0. The results are dramatically different, and even a high degree of adoption of the highest efficacy masks does not completely stop transmission. Note that even if the susceptible population don masks at a 100% rate, the mask wearing rates of detected asymptomatic and infected people are fixed at 90% (for those > 65) and 70% (for those younger) in the simulation, which helps explain the seemingly low performance of 100% mask wearing rate for N95 masks.

We estimated the incidence rate ratios (IRR) for new infections among mask wearers relative to non-mask wearers over the course of 6 months, for different types of mask (Table 2). These estimates reflect the impact of mask wearing on an individual wearer, whereas all of the other analyses in this paper are focused on the population-level impact. The IRR at a given point in time is the ratio of the number of new infections per capita among the mask wearing population to the corresponding number among the non-mask wearing population. This assumes equal mixing of masked and non-masked individuals – modeling the tendency for those populations to self-segregate would tend to decrease these IRR values. As expected, the greater the mask efficacy, the greater the difference in new infection rates as measured by the IRR. After 6 months, new infections are projected to occur at around half the rate among mask wearers compared to those not wearing N95 respirators, whereas in a scenario where medical procedure masks are worn, infections among mask wearers occur at around a 32% lower rate. We evaluated the impact of face mask usage rates and efficacy parameters on the effective reproduction number for R_0_ = 2.5 and R_0_ = 6.0, to represent the impact of highly contagious variants of concern (e.g., B.1.617.2) (Figure 4) [24]. Note that warmer colors corresponding to higher effective reproduction numbers are visible in the lower left-hand corner of the right panel but less so in the left panel. As we approach 100% source control and wearer protection efficiencies, masks reduce effective reproduction number < 1 for the low R_0_ scenario, but not for the high R_0_ scenario, given the same wearing percentages used to generate Figure 2. For example, when the baseline R_0_ = 2.5, an effective reproduction number of 1 is achieved by a hypothetical mask with source control and wearer protection efficacies of 84% and 60%, respectively. However, these same efficacies would result in an effective reproduction number of 2.33 when the baseline R_0_ = 6.0, as is likely the case with the Delta variant of concern. Those efficacies for masks are achievable with common cloth masks and medical procedure masks if they are doubled up, if the cloth masks have filter inserts, or if either type of mask is overfit with a fitter or brace to ensure a tighter fit [29-31]. If source control efficacy is 96% and wearer protection efficacy is > 70% (in line with efficacies for properly worn N95 respirators) then the effective reproduction numbers < 1.0 (R_0_ = 2.5) and = 2.19 (R_0_ = 6.0). Similarly, adoption of medical procedure masks (source control efficacy 56%, wearer protection efficacy 28%), results in effective reproduction numbers of 1.30 (R_0_ = 2.5) and 2.98 (R_0_ = 6.0). Please note that in Figure 4, even when source control and wearer protection efficacies of masks are zero, there is still some small measure of containment due to the reduced contact rates of those who are detected and infected (whether symptomatic or asymptomatic) in the simulation.

**Table 2:**
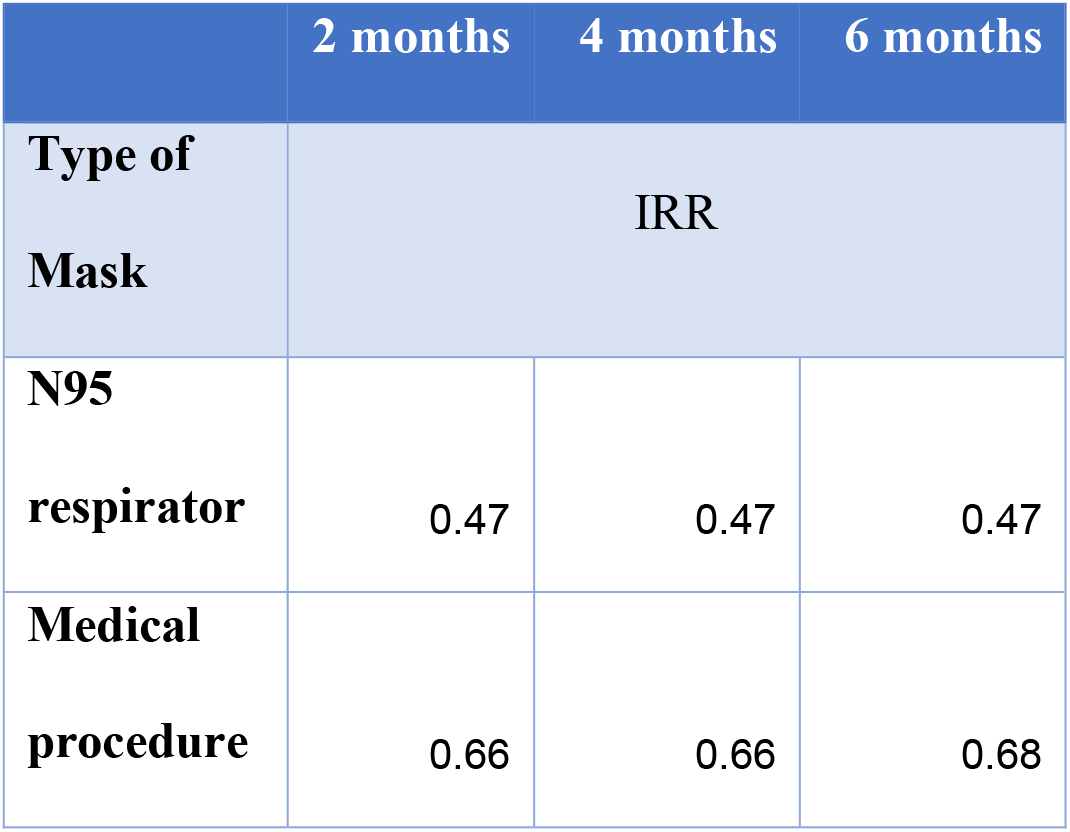
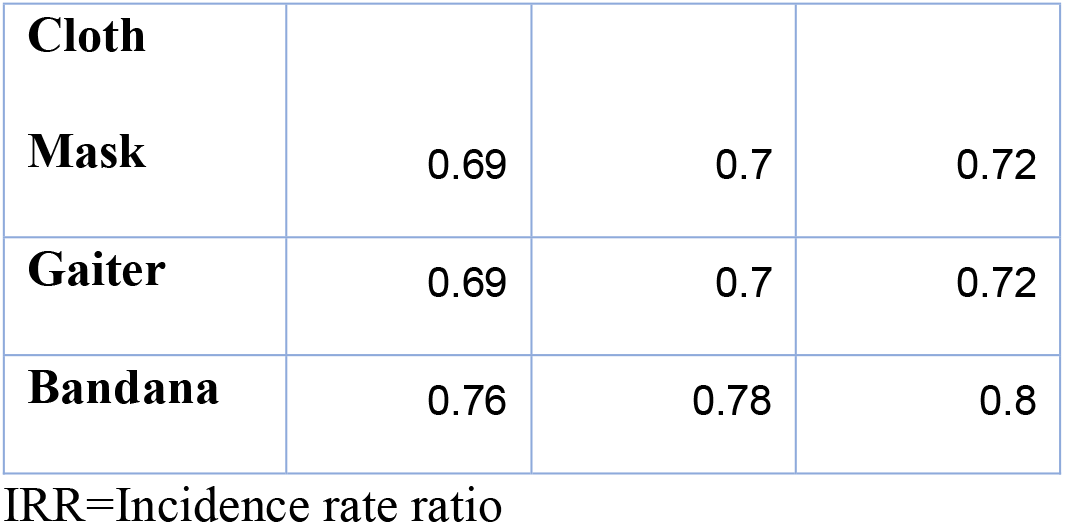
Incidence rate ratios (IRR) at 2-month intervals of new infections among masked vs. non-masked population. Each row represents a scenario in which all mask-wearing individuals are assumed to wear the specified type of mask. Wearer protection efficacy is assumed to be half of source control efficacy. It assumes 60% of the susceptible population <65 years old are wearing masks, 80% of those ≥65 years old wear masks, and both rates increase 10% for both detected and infected persons (whether symptomatic or asymptomatic).

**Figure 4:**
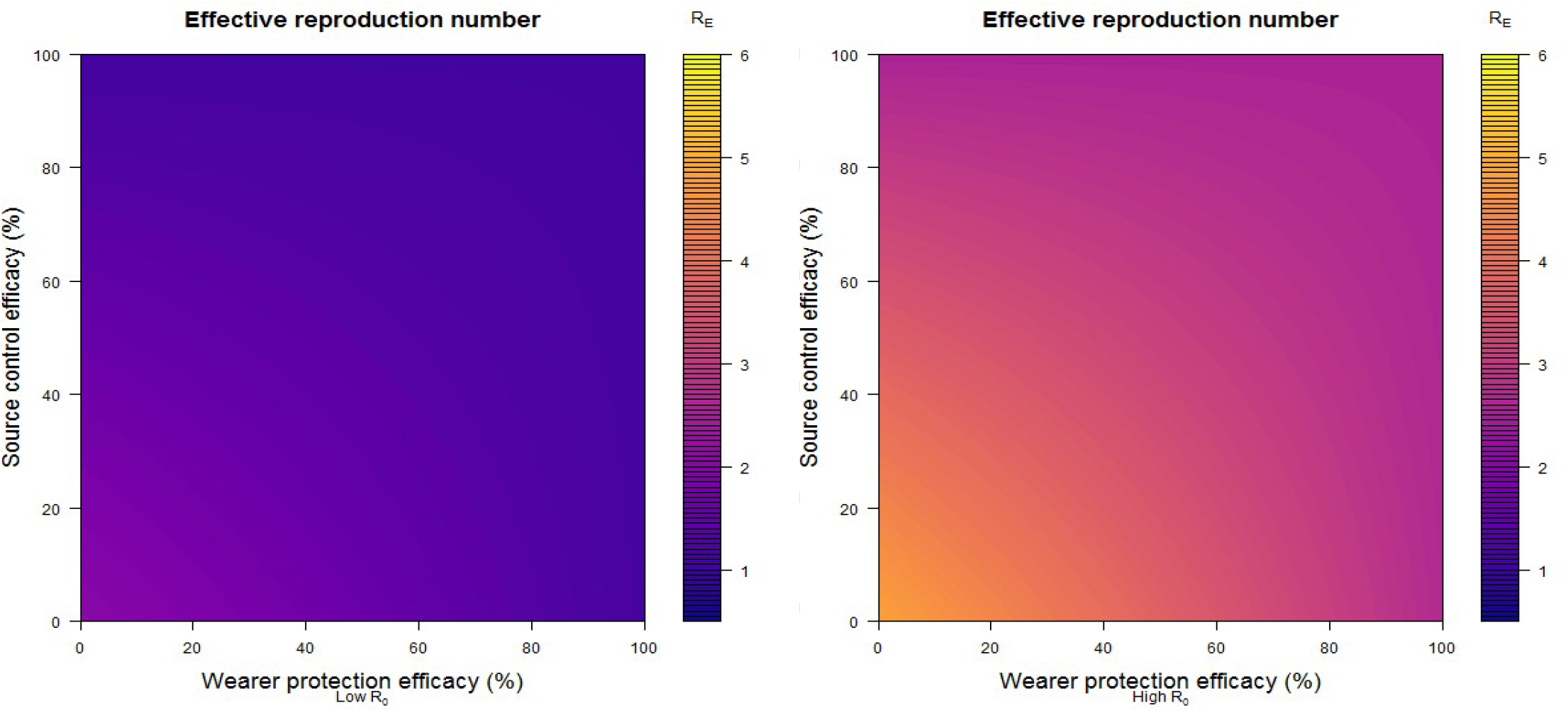
Effective reproduction number for given mask use by varying efficacy parameters shown on the horizontal and vertical axes. This analysis assumes 90% and 70% mask use rates for infectious and detected persons older and younger than 65 years of age, respectively, and 80% and 60% among susceptible persons for the same age breakdown. Asymptomatic detection and symptomatic awareness fractions are given in Table 1. The baseline R_0_ in the absence of mask use is assumed to be 2.5 in the left panel and 6.0 in the right panel.

## Discussion

Our results highlight the potential for substantial reduction in SARS-CoV-2 transmission, even with moderately effective masks, when they are worn consistently correctly (over the chin and covering nose and mouth) and/or per manufacturers’ specifications by a large portion of the population. These findings underscore the potential impact of population-wide measures that can control transmission from infected individuals who do not have symptoms, both pre-symptomatic individuals who are infectious prior to developing symptoms and individuals who never experience symptoms. By extending the Worby and Chang model, we evaluated the impact of different face mask use by age and highlight the need for wide adoption of these interventions. Pairing this modeling framework with laboratory-derived parameters for source control efficacy of different types of face masks helps to more accurately compare the relative efficacy of each mask type as an intervention. Even with more specific source control parameterization, the results are generally consistent with previous modeling studies [10, 11]: face masks with realistic source control efficacy can reduce transmission substantially, and widespread adoption can mitigate transmission at the population level. Furthermore, if the most common types of face mask – cloth and medical procedure masks – can be enhanced with more recent recommendations to improve fit around the nose and mouth, such as braces, elastic fitters, or even double masking, those substantial reductions can be improved upon.

Our study and several others suggest that the magnitude of reduction in SARS-CoV-2 transmission increases non-linearly with increased mask usage. The reasons for the non-linear multiplier effect are several, at least including potential epidemiological, immunological, and behavioral mechanisms [17, 27, 31, 32]. Non-linear terms are inherent in the mathematical mechanism of transmission reduction, given that masks act as both source control on the infected and personal protection on the susceptible, terms which are multiplied together in the transmission equations. This can be seen in the curvature of the line graphs of Figure 3 as mask usage increases (diminishing returns can be seen as mask usage increases towards 100% in Figure 3A for the N95 respirators, however). Furthermore, it is hypothesized that there are non-linear effects inherent in the pathogenesis of SARS-CoV-2 infection, in that masks reduce the initial viral exposure even if a wearer becomes infected despite the mask, decreasing the severity of infection, reducing viral load and shedding, and increasing the asymptomatic ratio [17, 32, 33]. If this hypothesis is substantiated and we ignore complications arising from a higher asymptomatic rate (i.e., more challenges with case identification), then there are potentially several non-linear terms describing how the reproduction number decreases with mask efficacy and use. Lastly, analysis of data on behavioral correlates of face mask use shows that people wear face masks more often when they see others do so, even when they already intended to wear a mask [28]. If changes in behavior were modeled, this would add another favorable non-linear term to the impact of mask wearing.

The pandemic literature does contain a minority of reports that do not confirm the efficacy of masks, although these studies have some important limitations. In particular, commentaries have been written about the methodological limitations of a recent publication by Bundgaard et al. that appears to question the efficacy of face masks [34]. [35, 36]. Specifically, the study was only powered to test if the wearer protection efficacy of medical procedure masks (referred to as “surgical masks” in Bundgaard et al.) was >50% and was not designed to measure their effect as source control (because it was estimated only 5% of the population were wearing masks at the time of the study). The Bundgaard et al. results were underpowered to detect wearer protection efficacies of medical procedure and cloth masks. This is similar to another randomized controlled trial (RCT) of cloth face masks as wearer protection against influenza virus infection among healthcare workers by MacIntyre et al. [2]: the study was designed to evaluate only the wearer protection effectiveness, not the source control effectiveness. Critically, the MacIntyre et al. study did not compare cloth masks to no mask, only to masks of the health workers’ choosing, potentially including medical procedure masks. Hence, this RCT *in a healthcare setting* did not have the negative control of not wearing a mask to help inform definitive conclusions about the efficacy of cloth face masks for the general population in non-healthcare settings. In fact, a follow-up study by MacIntyre et al. in 2020 found that healthcare workers whose cloth masks were laundered by the hospital were protected as well as those who wore medical masks [37]. Also, recent results from an epidemiological study [38] analyzing population level mask mandates where masks are more widely used are much more positive regarding the effectiveness of masks.

## Limitations

Despite widespread usage of masks and other mitigation strategies [39], transmission of SARS-CoV-2 remains inadequately controlled in the United States. There are many potential reasons why surveillance data and ecologic field studies might not show the magnitude of reduction in infections due to increasing mask usage predicted here. The parameters used in the models developed here might need to be better calibrated to match local transmission probabilities when individuals contact one another (either through direct physical contact, e.g. through a handshake or kiss, or a proximal, two-way conversation consisting of 3 or more words). Also, surveys indicating mask usage in the population may have overestimated adherence over time or the proper use or maintenance of masks. We model mask use as a set of parameters that can vary by age, but not by other societal subgroups, and our age groups were only divided into ≥65 years and <65 years. Furthermore, our model does not distinguish between differing contact rates within relevant populations such as schools, workplace, and households, but instead uses U.S.-national estimates for contact rates.

The source data for mask efficacy used in these models were derived from controlled laboratory simulations and not from human experiments. Measurements by other groups of filtration efficiency using actual human volunteers tend to show more variation, and in some cases the efficacies are lower than those reported here [40, 41].

Other limitations of the study are that mask usage is not assumed to vary over time, although it is likely that consistent and correct mask use may increase or decrease over time as individuals change their behaviors. Thus, we model homogeneous and unchanging mask use in a limited number of subgroups vs. the reality that mask wearing is heterogeneous according to mask type, sub-population, maintenance and proper use, and many other time-varying characteristics. This may result in over-estimation of the impact of face masks on the pandemic. If so, even higher mask uptake would be necessary to achieve substantial reductions in cases than is indicated here. Although the post-holiday 2020-2021 surge in cases seems large given a fairly high rate of mask usage, we have no solid counterfactual information for comparison [12], i.e. we do not know what the results would have been with no mask usage.

## Conclusions

Modeling studies, including this analysis, have estimated how face masks can reduce transmission of SARS-CoV-2 and make a major impact at the population level, even with varying levels of adherence and effectiveness of masks. Multiple public health interventions are needed to reduce the transmission of SARS-CoV-2 and, as our analysis shows, robust use of face masks is an important contributor. Face masks of various materials have the potential to substantially reduce transmission in the SARS-CoV-2 pandemic, depending on the type and fit of mask and the percentage adoption in the population. Furthermore, by attempting a more exact quantitation of the impact of masking, studies like this can show, for example, that for highly contagious new variants, such as the Delta variant of concern, masks alone are not enough to contain the outbreak, and other control strategies are needed (e.g. social distancing, hand washing, and vaccination). Public outreach and policies encouraging mask wearing, especially highly efficacious masks, need to be encouraged along with other prevention strategies. In fact, this study suggests that the current, imperfect use of masks has likely already reduced both cases and deaths.

## Data Availability

All data/parameters used in the models are reported in the manuscript. Code is available upon request.

## Disclaimer

The findings and conclusions in this report are those of the authors and do not necessarily represent the official position of the Centers for Disease Control and Prevention (CDC).

## Declarations

### Ethics approval and consent to participate

No human subjects were used in the study, therefore no consent was needed.

### Consent for publication

All authors consent to the publication. The paper has been cleared for publication by the CDC. The pre-print version of this article is present on https://www.medrxiv.org/content/10.1101/2021.04.21.21255889v1.

This article is not published nor is under publication elsewhere.

### Availability of data and material

R code is available upon request.

### Competing interests

All authors declare no competing interests.

### Funding

All authors are employees of the CDC – no external funding was needed.

## Authors’ contributions

BMG, ANH, and RBS conceived the study. BMG and ANH did the background research and wrote the primary draft of the manuscript. ANH served as the primary statistician for the project, adapted and modified the models and methods, and programmed the models and equations in R. BMG and Prabasaj Paul served as secondary statisticians and provided critical review of the models and equations, and BMG provided some ancillary models and equations. BMG and ANH contributed equally to the paper. All authors helped pull together the parameters needed for the models from the primary literature and other sources, and all authors contributed to writing and critical review of the final manuscript.

## Acknowledgements

The authors would like to thank all reviewers at the CDC who added to the clarity of the final manuscript, as well as other members of the CDC COVID-19 Response Modeling and Analytics Task Force for helpful discussions.

## Supplement

### Methods

Transmission of SARS-CoV-2 is modeled by a system of ordinary differential equations (ODE) and compartments corresponding to age, disease status, and mask-wearing status. Compartments comprise susceptible (*S*), exposed (*E*), pre-symptomatic (*P*), asymptomatic and undetected (*A*_*u*_), asymptomatic and detected (*A*_*d*_), symptomatic (*I*), recovered (*R*), and deceased (*D*). The total population is *N* = *S* + *E* + *P* + *A*_*u*_ + *A*_*d*_ + *I* + *R*. Initially, *N* = 100,000. Each of these compartments is further stratified by age (16 age groups from the POLYMOD study) and mask-wearing status (yes/no). Thus, each disease compartment is represented by a 16 *×* 2 matrix, with entries corresponding to the number of individuals of that particular disease status in age group *i* = 1,*…*,16 and with mask status *j* = 0 (no mask), 1 (mask). In matrix form, the ODE system is:

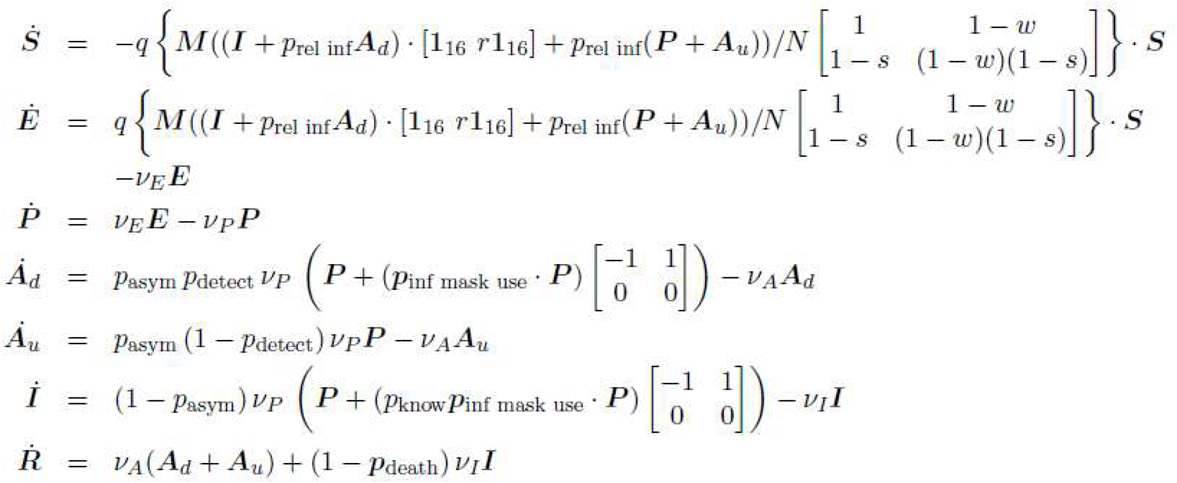

Dot superscript denotes derivative with respect to time; central dot *·* indicates pointwise multiplication of matrices of the same dimension, or of the columns of a matrix by a vector of the same dimension.

Compartment durations are specified by a rate *ν*_*J*_, where *J* is the compartment. Average duration in a compartment is 1*/ν*_*J*_. These rates model the durations of days exposed (2 days), pre-symptomatic (4 days), asymptomatic (9 days), and symptomatic (9 days). Relative infectiousness of pre-symptomatic and asymptomatic persons compared to symptomatic persons is *p*_rel inf_ = 0.75. Detection probability of an asymptomatic case is *p*_detect_ = 0.05 and the proportion of asymptomatic infections is *p*_asym_ = 0.30. The risk of death for symptomatic individuals was inferred from age-specific infection fatality ratios (IFR) via the equation CFR = IFR*/*(1 *− p*_asym_), where CFR denotes the case fatality ratio. IFRs are 0.00003 for ages 0-19 years, 0.0002 ages 20-49, 0.005 ages 50-69, 0.054 ages 70 and older. These parameters are based on the September 2020 estimates included in the CDC Pandemic Planning Scenario #5 [23]. We further assume that 20% of symptomatic individuals know that that they are sick with SARS-CoV-2 and we denote this fraction by *p*_know_. These people put on a mask.

POLYMOD daily contact rates were obtained from the study by Prem *et al*. [21]. The raw matrix of contact rates was adjusted in the usual fashion to maintain balance (numbers of contacts of age group *i* with age group *j* same as that of *j* with *i*). The raw matrix *C* is transformed to C’ by

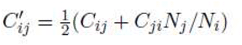

where *N*_*i*_ is the population size of age group *i*. Age distribution of the population was based on American Community Survey (ACS) estimates for the U.S. population [42]. Matrix C’ is, in turn, transformed to give the symmetric matrix *M* in the ODEs by

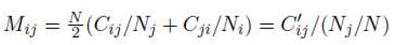

In this formulation, a typical term in the force of infection (FoI) arising from an age-stratified infectious compartment *J* = *P, A*_*d*_, *A*_*u*_, *I* of a given mask status is

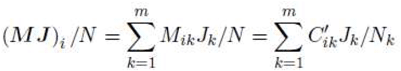

where *m* = 16 is the number of POLYMOD age groups corresponding to 5-year age bands ranging from younger than 5 years old to 70-75 years old, with the oldest age group comprising persons 75 years and older. The FoI is further stratified in the ODEs above depending on mask status. This is implemented by the 2 *×* 2 matrix appearing in the differential equations for *S* and *E*. This matrix governs reductions in the FoI according to source control efficacy (sce, or *s* for brevity) and wearer protection efficiency (wpe, or *w*) conferred by the mask type. These combine in four ways depending on the mask status of the infector and infectee (e.g., 1 *–* wpe and 1 *−* sce multiply together in the case of transmission by mask wearers to mask wearers). We further assume that symptomatic and detected, asymptomatic people who wear a mask have a lower, daily rate of contact. As contact rates in compartmental models apply to susceptible, not infectious, individuals, we model this as a reduction in infectiousness by a proportion *r* for both symptomatic and detected, asymptomatic mask-wearers. Specifically, if subscript *i* indexes age group and superscripts ‘none’ and ‘mask’ denote no mask and mask wearing, respectively, expanding the matrix formulation of the ODEs for susceptible individuals gives

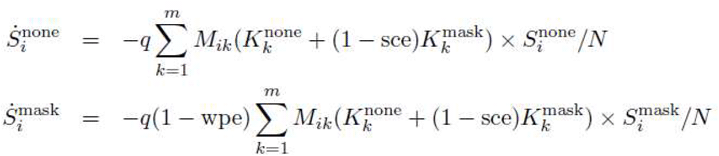

where 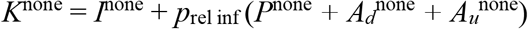 and

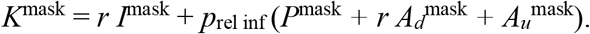

The age-specific vector *p*_inf mask use_ (in the set of 7 ODEs specified at the beginning of this supplement) specifies the proportion of symptomatic and detected asymptomatic people who don a mask upon learning they are infectious. The corresponding proportion for symptomatic individuals is *p*_know_ × *p*_inf mask use_. The 2 *×* 2 matrix appearing in the differential equations for *A*_*d*_ and *I* governs the adoption of masks by asymptomatic individuals when they are detected (the first column corresponds to no mask, the second to mask wearing). In simulations, we assumed that individuals aged 65 years and older adopted masks at one proportion and younger than 65 years at another, lower, proportion (but this can be changed by the user). Expanding the matrix formulation of the ODEs for detected asymptomatic individuals gives

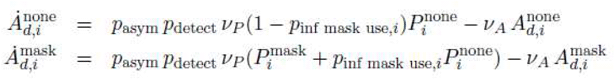

It is assumed that a proportion of the general population (susceptible individuals) wear a mask at the outset and keep it on at all times (or at least when mixing in the population). This proportion can vary by age. We assume that in the general population, 80% of those aged 65 and older and 60% of the rest, wear a mask (and keep it on indefinitely).

To seed the epidemic, we arbitrarily assumed that there were 10 detected, asymptomatic non-masked individuals in each age group at the outset. Time units were expressed in days. The model was run for 6 months (183 days) with a timestep of 0.25 days using a Runge-Kutta solver in R v.3.6.3 [43] using the package ‘deSolve’ [44].

The FoI was calibrated to yield a basic reproduction number *R*_0_ = 2.5 for the sub-model without mask usage. This yielded the parameter *q* = 0.01429, which represents the probability of a symptomatic infectious person infecting a susceptible person upon contact between them. The reproduction number was calculated as the dominant eigenvalue of the next-generation matrix (NGM) using the method of van den Driessche and Watmough [45]. Computation was facilitated by the R package ‘blockmatrix’ [46] owing to the sparseness of the matrices involved. Details of this calculation are described further below.

### Calculation of R_0_

Following [45], we construct matrices *F*, describing rates at which infectious individuals produce new infections, and *V*, consisting of all other rates, whose inverse describes average durations in compartments. The *i*th row and *j*th column of these matrices is the partial derivative of the right-hand side of the differential equation for compartment *i* with respect to compartment *j*, evaluated at the disease-free equilibrium (DFE). Only the 5 infected compartment types are considered, namely, *E, P, A*_*d*_, *A*_*u*_, *I*, enumerated by age group and mask status. The basic reproduction number *R*_0_ is given by the dominant eigenvalue of the NGM *F V* ^*−*1^.

Matrices *F* and *V* are of dimension 160 *×* 160 (2 mask statuses *×* 16 age groups *×* 5 relevant compartment types). However, as new infections only arise from the *E* compartments, via the previously described FoI, matrix *F* is sparse. So too is *V*, as a lot of its sub-blocks are zero or diagonal matrices. Hence, we can construct these matrices in block form. We use the Kronecker product of matrices, which we denote by ⊗.

The DFE depends on the initial age-specific proportions, *p*_susc mask use_, of the general, susceptible population wearing a mask. Multiplying the ACS age group proportions pointwise by *p*_susc mask use_ and 1 *− p*_susc mask use_ gives the age-specific proportions of the population with and without masks, respectively. These are multiplied by the hypothetical total population size *N* = 100,000 to obtain numbers in each stratum.

Vectorizing the 16 *×* 2 matrices in the ODEs by stacking columns into a single 32 *×* 1 column vector, we have the following constituent matrices for calculating *R*_0_:

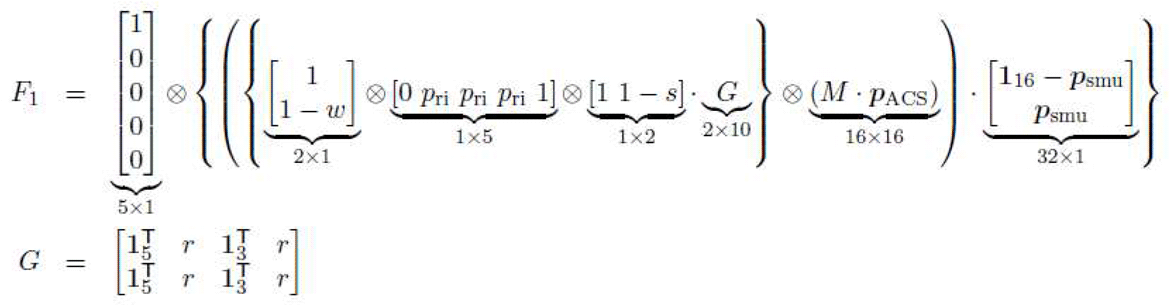

For brevity, *p*_r.i._ denotes *p*_rel inf_ above, *p*_ACS_ is the ACS population age distribution, *p*_s.m.u._ denotes *p*_susc mask use_ above, w denotes WPE, s denotes SCE, **1**_*n*_ is the *n*×1 vector of 1’s, and *·* represents pointwise multiplication by column. Matrix *M* is the POLYMOD-derived contact matrix described earlier. It follows from the definition of Kronecker product that *F*_1_ is 160 *×* 160. The order of entries in row or column vectors of length 5 corresponds to the compartment types *E, P, A*_*d*_, *A*_*u*_, *I*. Thus, the 5 *×* 1 vector on the left represents new infections only arising from compartments of type *E* (component 1) and not from *P, A*_*d*_, *A*_*u*_, *I* (components 2 to 5) and its occurrence renders *F*_1_ sparse. The 1 *×* 5 row vector has components 2-4 as *p*_rel inf_, indicating the relative infectiousness of presymptomatic and detected and undetected asymptomatic individuals (*P, A*_*d*_, *A*_*u*_) compared to symptomatic individuals *I* (component 5). Row and column vectors of length 2 correspond to mask efficacies. The vector *p*_ACS_ represents the population age-distribution and the 32 *×* 1 vector on the right denotes the age-specific general population mask wearing proportions.

The matrix *F* is given by *F* = *qF*_1_, where *q* is the calibration parameter representing the probability of symptomatic infectious persons infecting susceptible persons upon contact between them.

Matrix *V* may also be expressed in block form as:

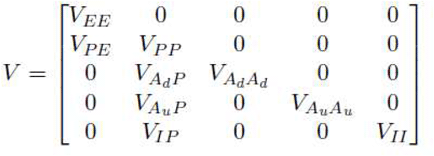

where each block is a 32 *×* 32 matrix. Theory guarantees *V* is invertible. As with *F*, the order of the component types in block rows and columns here is *E, P, A*_*d*_, *A*_*u*_, *I*. The nonzero blocks are as follows, with parameter notation as given earlier, and *I*_*n*_ denoting the *n × n* identity matrix:

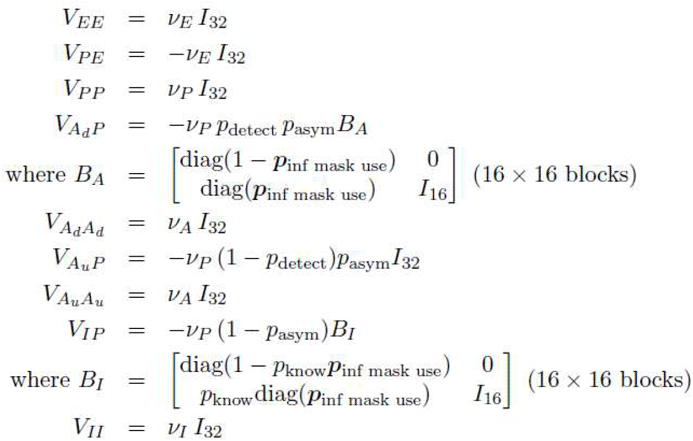

Blocks *V*_*XX*_ on the leading diagonal correspond to outflows from compartment type *X*. Off-diagonal blocks *V*_*XY*_ correspond to inflows from compartment type *Y* to compartment type *X*. The *B* matrices correspond to the adoption of mask use (change of mask status) upon asymptomatic detection/symptomatic awareness, according to the age-specific proportion (*p*_inf mask use_) who do so. Denoting the dominant eigenvalue of a matrix by *ρ*, we have

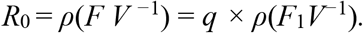

Setting baseline *R*_0_ = 2.5 (no mask use), we calibrate *q* = 2.5 *÷ ρ*(*F*_1_*V*^*−*1^).

### Computation of symptomatic and asymptomatic detection rates

The percentage of symptomatic and asymptomatic SARS-CoV-2 cases that are detected are not established numbers, but they are suspected to be low given the general detection rate of 16.1% [47, 48]. Both can be estimated with a simple Bayesian calculation, however, given the general detection rate (P(case) in the equation below), the asymptomatic rate of infections of 0.3 [23], and the probability that detected cases are and remain asymptomatic (0.2) or symptomatic (0.8) [49]. For example, the probability that a person becomes a detected case (e.g. through contact tracing efforts) given the person is asymptomatic is given below

P(case|asymptomatic) = P(asymptomatic|case) P(case)/P(asymptomatic)

The simple calculation yields a 10.7% detection rate for asymptomatic individuals, and a 18.3% detection rate for those with symptoms. The 18.3% figure appears to be in line with epidemiological estimates as well [50].

## Notes

### Competing Interest Statement

The authors have declared no competing interest.

### Clinical Trial

This is an epidemiological modeling study, not a clinical trial

### Funding Statement

No external funding was received.

### Author Declarations

No IRB was needed as this is an epidemiological modeling study.

### Summary of Updates

The manuscript has been updated to including modeling for the impact of the delta (B.1.617.2) variant on viral control using face masks.

